# Investigating dynamics of COVID-19 spread and containment with agent-based modeling

**DOI:** 10.1101/2020.08.18.20177451

**Authors:** Amirarsalan Rajabi, Alexander V. Mantzaris, Ece C. Mutlu, Ozlem O. Garibay

## Abstract

Governments, policy makers and officials around the globe are trying to mitigate the effects and progress of the COVID-19 pandemic by making decisions which will save the most lives and impose the least costs. Making these decisions needs a comprehensive understanding about the dynamics by which the disease spreads. In this work, we propose an epidemic agent-based model that simulates the spread of the disease. We show that the model is able to generate an important aspect of the pandemic: multiple waves of infection. A key point in the model description is the aspect of ‘fear’ which can govern how agents behave under different conditions. We also show that the model provides an appropriate test-bed to apply different containment strategies and this work presents the results of applying two such strategies: testing, contact tracing, and travel restriction. The results show that while both strategies could result in flattening the epidemic curve and significantly reduce the maximum number of infected individuals; testing should be applied along with tracing previous contacts of the tested individuals to be effective. The results show how the curve is flattened with testing partnered with contact tracing, and the imposition of travel restrictions.

## 1 Introduction

COVID-19 has posed a distinct challenge to the world, the likes of which have been rarely witnessed before. It is yet another historical example for the concern that a more connected and intertwined world, has brought us more stability and fragility at the same time [37]. The recent catastrophe of COVID-19, has swept through all continents and most countries [36]. As of the time of writing this manuscript, 18.7 million cases have been reported, with a death toll in the hundreds of thousands [30]. In the absence of a vaccine or a definite treatment for the disease, governments and officials have enforced isolation rules to control the increasing number of infected people and take off the burden put on healthcare systems and workers.

Understanding the dynamics by which the disease spreads and making the right decisions is vital in combating and containment of this disease. For more than a hundred years, mathematicians and later epidemiologists have put effort into making models that can predict statistical properties of epidemics with focus on key parameters like the *R*_0_ [17]. Besides predicting the future state of a pandemic and the number of infections involved, a model to assist decision and policy makers to take the necessary and optimal decisions is a necessity. In the case of COVID-19, some governments have adopted epidemiological models and enacted urgent poilicies based on those models (see for example [27, 42]).

Epidemiological models can be divided into two groups, that of the *deterministic* and the *stochastic*. There is the classical Kermack Mckendrick model(SIR) [21], SI [25], SEIR [26] which are all classified as deterministic models and their features are discussed by [31]. The other group of epidemic models, the deterministic, mostly draw on the theory of Markov processes [9] which are generally more reliable when dealing with populations of smaller sizes.

While providing insight into the threshold nature of epidemics [11], the majority of these traditional models do not incorporate direct contact between individuals as they treat individuals as an aggregate collection. They make demanding unrealistic assumptions such as perfect mixing, usually missing the heterogeneity of agents’ behaviors, which could have nonlinear effects (e.g. super spreaders from occupations that frequently travel). They also often fail to address complex spatial and temporal factors such as variable population densities and dynamics.

Agent-based modeling approaches allow for a detailed focus on the dynamics of interactions between individuals [20]. They can be used to capture complex social networks and the direct contacts between agents and can overcome the limitations of compartmental and meta-population models [15]. These models have been successfully applied to simulate the spread of 1918 “Spanish Flu” epidemic [2, 10, 28], spread of cholera [5], seasonal influenza outbreaks [15], spread of H1N1 [22], and Ebola epidemic [34]. Besides being able to incorporate links contained in individual mobility data, agent-based modeling approaches can explore and investigate the complexities of transmissions of diseases over space and time. They can be integrated with geographic information systems (GIS) and account for the geospatial context of the spread of communicable diseases [29]. The work of [6] is an example of utilizing crowd-sourced geographic information to develop an agent-based model that simulates the aftermath of a catastrophic event. Other studies have also attempted to employ geographic information systems to build spatially explicit agent-based models and simulate and investigate the spread of diseases [8, 43].

Multiple computational studies are being proposed to tackle COVID-19 pandemic. The work of [40] has proposed a coupled SIR with opinion dynamics epidemic model which evaluates the effect of opinions on severity of an epidemic. [3] developed an agent-based model that simulates the spread of COVID-19 in Australia and applies several intervention strategies and compares the results. In [14] the authors proposed a new SIR type model in which the disease reproduction number evolves dynamically with time to account for societal and state official reactions. The work of [7] provides an agent-based model of COVID-19 to evaluate the transmission risk in facilities, by considering individual profiles for agents that defines their social characteristics. The work of [35] proposes an agent-based implementation of SEIR model that involves people, businesses and government, calibrates it for the case of Brazil, and applies seven social distances interventions and reports the results. [1] develops a network epidemic model and investigate a threshold that instigates the public awareness and its effect of the final size of the epidemic.

The rapid spread of COVID-19 has forced many governments to apply various interventions and containment strategies their respective societies (eg that in Italy [13]). There are a range of common intervention strategies being applied by governments, such as applying travel restrictions, testing and contact tracing, school closings, preventing public gatherings, and others [16]. While these interventions are vital in combating the spread of the disease until a vaccine or a definite cure is secured, there are unavoidable consequences, primarily economical. Therefore, it’s imperative to analyze the effectiveness and consequences of different containment strategies. On the other hand, while numerous epidemiological modeling efforts are put forth to investigate the size and effects of COVID-19 pandemic and under different intervention strategies, little attention has been paid to developing models that account for the possibility of multiple waves of the disease spread. In this study, drawing on the work of [12], we develop a variation of that model which also accounts for the incubation period of COVID-19. We also test two containment strategies on the model, present the results and show that the model could be used as a test-bed for experimenting various kinds of intervention strategies under different scenarios.

Epstein et. al. proposed a coupled contagion model of disease and fear dynamics [12]. Contagion of fear could be interpreted in the context of emotional contagion [23]. It should be noted that the fear variable in the work of Epstein et. al. is a more general term than of being ‘scared’, and rather should be interpreted as a ‘concerned awareness’ which is a ‘*behavior-inducing transmissible signal distinct from the pathogen itself*’ [12]. Such a factor can provide motivation for agents to engage in activities (such as self isolation) based upon their own understanding of the situation. One exceptional feature of this model is the ability to produce multiple waves of contagion, the phenomena which was observed in 1918 pandemic of “Spanish Flu” [19], and is currently occurring in multiple places during the COVID-19 pandemic. Being able to foresee these ‘waves’ is crucial in order to prepare in advance.

In this work we propose a model of contagion of disease and ‘concerned awareness’. Agents who interact with others could be infected by the disease, by fear, or by both. Section 2 explains the model in details; The two simultaneous and coupled transmission processes are explained, as well as the states that an agents can occupy and parameters of the model. We show that the model could also be implemented as a classical well-mixed ordinary differential equation, and provide the governing system of equations, as well as derive the required terms to calculate the basic reproductive number (*R*_0_). Section 3.1 describes the ability of the model to produce multiple waves of infection. Finally, Section 3.2 describes the containment strategies that are tested on the population of agents, along with presentation and interpretation of the results.

## 2 Methodology

### 2.1 Transmission of disease and fear

The model used in this study is inspired by the work of [12]. There are a total of 10 states that an agent can occupy in the model. The main four states of an agent in the model are as follows: 1) being a susceptible agent (S) that might get infected by fear or disease, 2) being an infected agent with disease (I) that is aware of the infection and has apparent symptoms, 3) an exposed agent (E) which is infected by the disease, is contagious, but doesn’t have apparent symptoms yet and is not aware of their own infection, and 4) a recovered agent (R) which is taken out of the cycle. The remaining six states that an agent can occupy are combinations of the ones mentioned. All of the ten possible states of the model is presented in Table 1 and discussed in Section 2.2.

**Table 1:** Possible states of agents in the epidemiological model

| state | description |
| --- | --- |
| S | Susceptible |
| E | Exposed |
| F | Feared |
| EF | Exposed and Feared |
| I | Infected |
| IF | Infected and Feared |
| QF | Self-quarantined Feared |
| QEF | Self-quarantined Exposed and Feared |
| QIF | Self-quarantined Infected and Feared |
| R | Recovered |

There are two contagion processes at play:

1. Transmission of *disease*: in this model the disease can be transmitted by an interaction with a disease-infected agent. This disease infected agent might be in their incubation period and show no symptoms, in which case we call them *exposed* shown with E, or be infected and their infection be obvious, in which case they are infected and shown with I.
2. Transmission of *fear*: Fear is transmitted between agents. Agents can contract fear by interacting with agents that are feared (F, IF, EF), or by interacting with agents that are infected and have symptoms (I). As discussed before, fear in this model should be looked at as a concerned awareness. This quality gives the model a more realistic essence, by giving agents an incentive to put themselves into self-isolation (In the absence of such as a contagion process and the absence of a feared population, the model reduces to a simple SEIR epidemic). In this model, agents that have isolated themselves because of fear, get out of self-isolation with a certain rate that is explained in Section 2.3.

### 2.2 Possible states of an agent

An agent can only belong to one of these states at any time. A *susceptible* agent (S) is one that is neither infected by the disease nor by the fear yet. A *feared* agent (F) is one that is infected by fear. An *infected* agent (I) is one that is infected by the disease and is aware and has symptoms. On the other hand, an agent which is infected and is contagious (i.e., can infect others) but shows no symptoms yet and is unaware of his infection, is in *exposed* state (E). The exposed state is imperative in modeling the dynamics of COVID-19, as the incubation period reported for this virus has reported to extend to 14 days, with a median of 5.1 days [24]. Contrary to assumption made in most implementations of SEIR mode, an exposed agent with state E is infectious and infects other agents with whom he interacts. In this model, an agent with *exposed* state always transforms to *infected* state after 5 time steps (representing 5 days).

An agent with an SE state is one that is both infected by fear and also exposed to infection (a yet incubated infection). State of IF shows an agent that is infected by fear and the disease (has symptoms and is aware of his infection). An agent with state of QF is one that is infected by fear and has isolated himself (quarantined). A state of QEF shows an agent which has underlying infection (exposed), infected by fear, and also has isolated himself from the rest of the population (quarantines). A state of QIF shows an agent that is infected by the disease and fear, and has put himself into isolation. Finally, a state of R shows an agent that is recovered from the disease and is taken out of the cycle. Table 1 shows all possible states that an agent can occupy during the course of the simulation.

### 2.3 Parameters

The model parameters are as follows:

- *α*: Per-contact fear transmission rate
- *β*: Per-contact disease transmission rate
- *λ*_1_: Rate of removal of those infected with fear (F, EF, and IF) to self-isolation (QF, QEF, and QIF, respectively)
- *λ*_2_: Rate of recovery (R) of all agents infected with disease (I, IF, and QIF)
- *ϵ*: Rate of progression from exposed to infected (the reciprocal is the incubation period).

Figure 1 shows all states, their relationship and transition rates between these states. The model subsumes the classic SEIR model, and it assumes constant population. Lets review some possible interactions between agents with different states and the outcome: a susceptible (S) that interacts with an exposed agent (E), remains susceptible with probability (1 *—β*) or turns into an exposed (E) with probability *β*. As another example, a susceptible (S) which interacts with an infected and feared agent (IF), stays susceptible with probability (1*— α*)(1*— β*), becomes exposed (E) with probability (1*— α*)*β*, becomes only feared with probability (1*— β*)*α*, and finally becomes both exposed and feared (EF) with probability *αβ*. Agents which are feared (F, EF, IF), go into self-isolation with rate *λ*_1_. Agents which are in self-isolation on the other hand (QF, QEF, QIF), return back to the cycle with rate *H*, and turn into S, E, and I, respectively. In the end, it should be noted that agents that are exposed (E, EF, QEF), turn into infected (I, IF, and QIF, respectively) after they pass their five incubation days.

**Figure 1:**
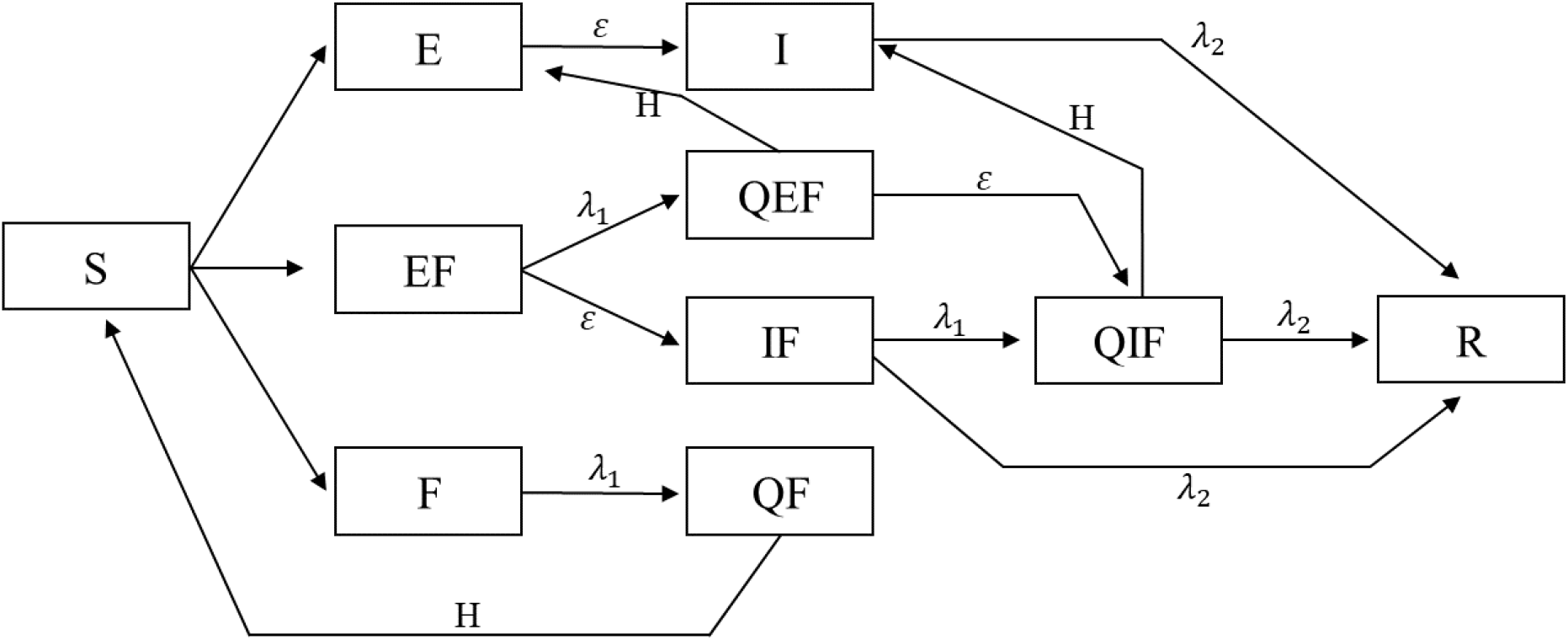
Model Flowchart. Here, all ten possible states of an agent in the model are depicted. In addition, parameters of *λ*_1_ shows the rate at which agents self-isolate due to fear, and parameter *λ*_2_ shows the rate at which disease infected agents recover from the disease. Parameter H shows the rate at which agents finish their self-isolation and return to circulation.

### 2.4 Differential equations and basic reproduction number

The proposed model can also be implemented as a classical well-mixed ordinary differential equation (ODE) system. Appendix Appendix 1 presents the generalizations and governing differential equations. The basic reproduction ratio, *R*_0_ is one classical epidemiological measure which is associated with the reproduction power of a disease. It is an important threshold quantity and denotes the *average number of secondary cases that are produced by one infected individual into a population of susceptible individuals* [41]. The details of calculating this number for the model is discussed in Appendix Appendix 2. The calculated number with respect to the parameter setting that was used for running the experiments was calculated to be 1.95.

## 3 Results

### 3.1 The *second wave* of infection

The case of 1918 pandemic of Spanish Flu has brought our attention to the concern of a possible *second wave* [44]. The proposed model is able to generate multiple waves of infection. Figure 2 shows two instance of this resurgence of infection cases. The mechanism by which the second wave is generated in Figure 2a is as follows: as the number of infections starts to decrease, individuals that had isolated themselves start exiting their self-isolation and returning back to the cycle. This phenomena is simulated by increasing parameter *H*, which is analogous to the case that individuals stop fearing about the disease and presume that epidemic is over and starting coming out of self quarantine. Looking at Figure 2, it should be noted that the second wave happens with a delay with respect to the sudden change of parameter *H*, which is manifested in the sudden fall in the number of quarantined agents. In Figure 2b on the other hand, the second wave is produced by a sudden fall in all *λ*_1_ parameter.

**Figure 2:**
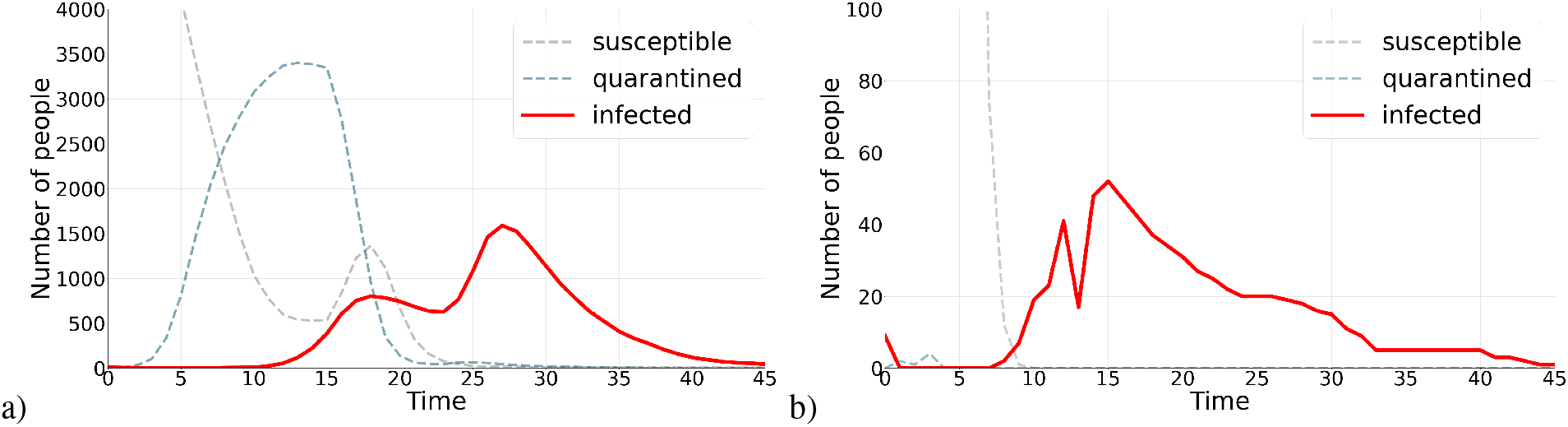
Two waves of infection. Figure a) shortly after the number of infected agents starts decreasing, quarantined agents start coming out of self isolation, a sudden increase in parameter *H*, which results in occurrence of a second wave of infections. Figure b) a sudden decrease in *λ*_1_ parameter, which show the rate at which agents in a state of fear quarantine themselves, produces a second wave of infection. These two scenarios describe how a second wave can be produced under different parameterizations.

### 3.2 Applying containment strategies to *flatten the curve*

As the number of COVID-19 cases surges, the term *flatten the curve* is very commonly used. The idea that a virus’ spread slows down so that fewer people need to seek treatment at any time is called ‘flattening the curve’. Flattening the curve is important so as to avoid the health care system from getting overloaded beyond its capacity and also averting the adverse economical impacts of the pandemic [38]. In this work we investigate two containment strategies that are mostly used by governments and officials: 1-testing and contact tracing 2-applying travel restriction and. In testing each scenario and model run, we record the maximum number of people that are infected with the disease and have symptoms: I, IF, and QIF. We call this recorded number *peak infection*; A flattened curve will have a lower peak infection number, as it has been flattened and its maximum must have become smaller.

The model is implemented in NetLogo [39]. The number of agents is kept 5000 in all simulations, and the world is a 160 × 160 grids of patches which form a two dimensional lattice. In each timestep, agents are first randomly positioned in patches, and then each agent looks at its neighbors (8 surrounding patches) and uniformly chooses one of its 8 neighbors to interact with. The result of the interaction between the two agents is determine by rules of interaction discussed in Section 2.3.

#### 3.2.1 Testing and contact tracing

While merely testing individuals will not stop the spread of the disease, WHO recommends a combination of testing and tracing the contacts of individuals that have been tested positive [33]. The successful containment of COVID-19 in some countries such as South Korea are attributed to their rigorous testing and contact tracing and isolation strategy [4]. In this section, we apply this testing and contact tracing strategy on the proposed model and investigate the results. The parameters used in this model are all fixed on 0.2, that is to say *λ*_1_ = *λ*_2_ = *H* = 0.2. Per-contact disease transmission rate(*β*) and per-contact fear transmission rate(*α*) are also fixed on 0.2. two quantities determine the experiment: *daily testing size* and *number of contacts tracing*. In each timestep, a number of agents, the number of which is determined by *daily testing size*, are chosen and tested for being infected with the disease. If they are tested positive, that is to say they have a state of E, EF, I, or IF, they are then put into isolation (QEF, or QIF), and their previous contacts are traced, tested, and put into isolation either if they are also positive. The number of their previous contacts which are traced, is determined by *number of contacts tracing* parameter.

Three daily testing size of 1, 100, and 200 were tested. Also, three number of contact tracing sized were tested: 1,10, and 20. Each experiment was run 2000 times (a total of 3 ×3 × 2000 = 18000 simulations) and the quantity that was recorded in each experiment, as explained in Section 3.2, is *peak infection*(the maximum number of people that are infected with the disease at any time during the course of the simulation).

Figure 3 shows the results of the simulation. For each parameter setting, peak infection number in all 2000 experiments are recorded and then probability density function (PDF) of peak infection is drawn. The results of the experiment show that, as the size of daily testing size increases, the median of peak infection also decreases (compare the median of 1774 when daily testing size and contact tracing numbers are 1, with the median of 1384 when daily testing size is 200 and number of contact tracing is set on 20). A Mann-Whitney U test between the two distributions (distribution 1: daily size = 200 and tracing size = 20 and distribution 2: daily size=1 and tracing size = 1) was conducted and a p-value of 2.28× 10^−123^ shows that the two distributions are significantly different. Perhaps, the most interesting result of this experiment is that the importance of contact tracing is revealed with large daily testing size. Increasing daily testing size, needs to be joined with contact tracing, in order to be really effective. This effect is pronounced with higher daily testing size numbers (Figure 3b, and Figure 3c). A Mann-Whitney U test between the two distributions (distribution 1: daily size = 200 and tracing size = 20 and distribution 2: daily size=200 and tracing size = 1) were conducted and a p-value of 1.24 × 10^−25^ shows that the difference between the two distributions is statistically significant.

**Figure 3:**
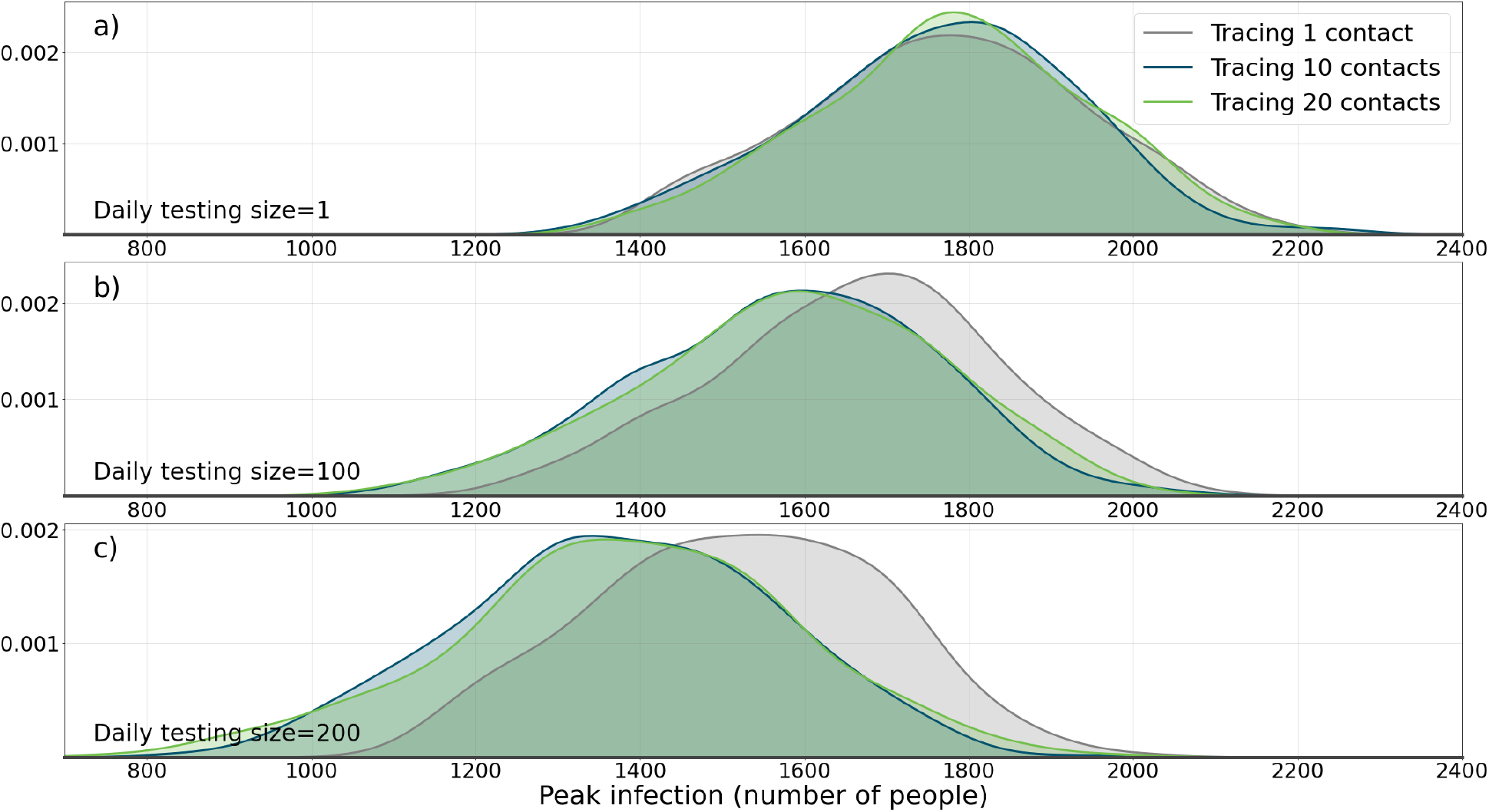
The effect of applying containment strategy I (testing and tracing) on the population. Here the effects of daily test size and contact tracing size on pdf (probability density function) of maximum daily number of infected agents during the course of epidemic is shown. a) A daily testing size of 1 is applied with three tracing sizes of 1, 10, and 20. b) Daily test size of 100, with tracing sizes of 1, 10, and 20. c) Daily test size of 200, with tracing sizes of 1, 10, and 20. It can be seen how testing and contact tracing together are required in order to change the distribution.

#### 3.2.2 Travel restriction

Travel restrictions and limitations are among the most common government interventions during the COVID-19 pandemic and it has been shown to be an effective measure [32]. In this section, we simulate application of this intervention in our proposed model and investigate its effect. This experiment could be analogous to travel restriction and human mobility restrictions set in place in some countries such as Italy in earlier days of the pandemic.

As mentioned in Section 3.2 the world in which the model is implemented is a 160× 160 grids of patches which form a two dimensional lattice. For this experiment, the world is divided into 4 quadrants: region I with positive x and y coordinates, region II with negative x coordinate and positive y coordinate, region III with negative x and y coordinates, and region IV with positive x coordinate and negative y coordinate. Upon starting the simulation, all 5000 agents are created and randomly put into a patch, which happens to be in one of the four regions. Contrary to the previous experiment, agents are only allowed to move and change positions within the boundaries in which they were created and are not allowed to change their region. For instance, an agent that is created in position (20,30) is not allowed to move to (−15,30) or (27,-48). The model is run for 4000 runs with no restrictions, and 4000 times with the restrictions present, and in each run the peak infection number is recorded. Similar to the previous experiment, all parameters are fixed at 0.2. The results of the simulation are shown in Figure 4. It is observed that by applying the travel restriction, the median of peak infection distribution is moved from 1775 to 1659. A Mann-Whitney U test between the two distributions was conducted and a p-value of 1.79× 10^−8^ shows that the two distributions are significantly different. A more pronounced effect could be reached by applying more extensive restrictions.

**Figure 4:**
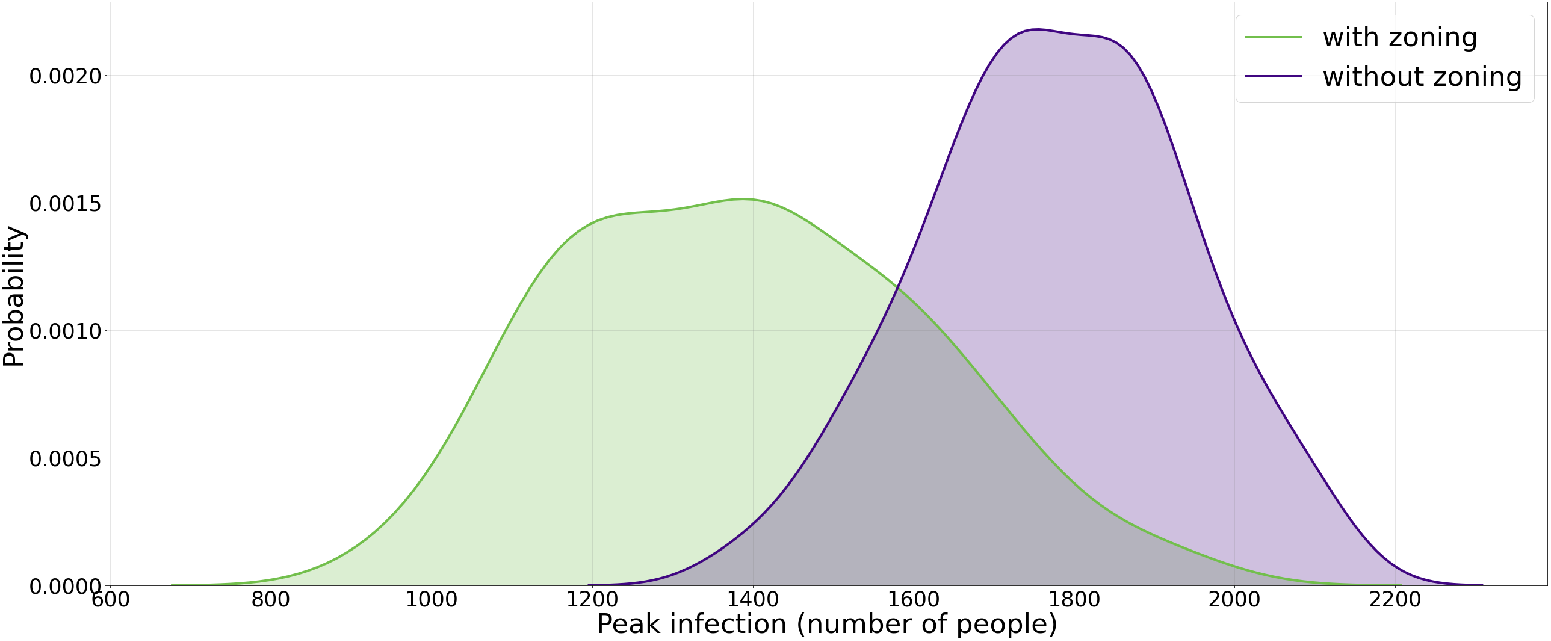
The effect of applying containment strategy II (zoning and travel restriction) on the population. This figure shows the effect of applying travel a restriction strategy on the p.d.f. (probability density function) of maximum daily number of infected agents during the course of epidemic. Here the zoning applies to agents so that they do not cross different areas of the urban space which is anchored from the initial position.

Figure 5 shows the effectiveness that applying both containment strategies have on *flattening the curve*. In Figure 5a, both simulation runs are performed with the same random number generator seed. As could be observed, test and contact tracing strategy is able to reduce the maximum number of infected agents by 27%. Figure 5 also shows two simulations with the same random number generator seed. The results show that applying travel restriction strategy is also effective in reducing the maximum number of infected agents by 31%.

**Figure 5:**
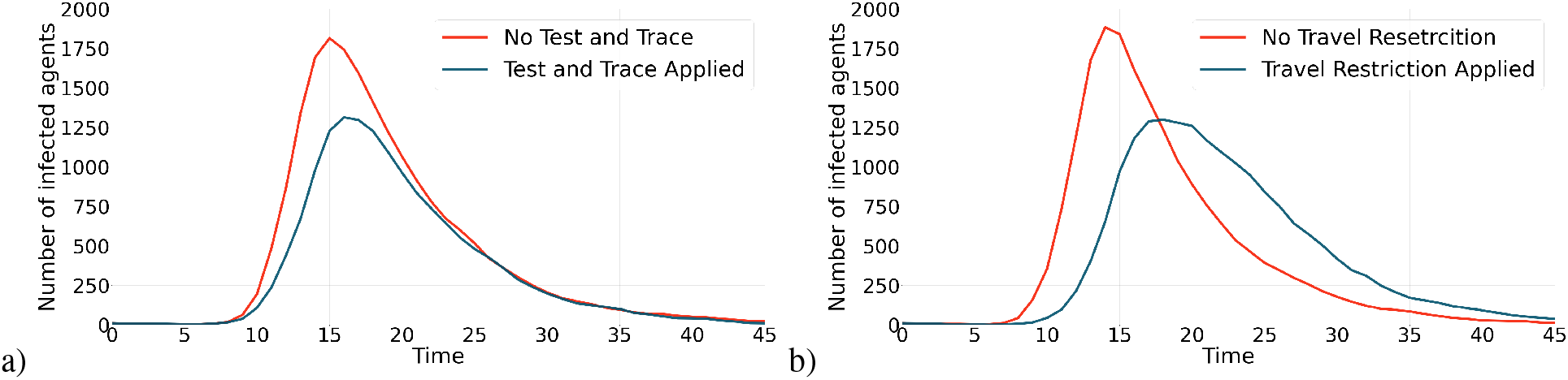
The effect of applying containment strategies (test and contact tracing and travel restriction) on the contagion curves. This figure shows the effect of applying the containment strategies on epidemic curves. a) Applying test and contact tracing has reduced the maximum number of infected agents by 27%. b) Applying travel restriction has reduced the maximum number of infected agents by 31%.

## 4 Conclusions

In this work, we provided an agent-based model of epidemics to simulate the spread of COVID-19 disease. The model is an SEIR model that is endowed with fear component that acts as an incentive for agents to self-isolate and get out of epidemic cycle. We showed that the model can also be implemented as a classical well-mixed and provided the process to calculate basic reproductive number of the disease which is simulated. One significance of this model is it being capable of producing multiple waves of infections which is imperative in studying dynamics of COVID-19 epidemic. We applied two common containment strategies that are practiced by governments to the model: 1) testing and contact tracing, and 2) travel restriction. The results of the experiments showed that both strategies could be effective in flattening the curve, i,e, reducing the maximum number of infected individuals in a single day (highest), over the course of the epidemic. In testing and contact tracing strategy, it’s important to note that the results of experiments show that testing alone is not adequate and should be applied along with contact tracing to be effective in flattening the curve.

A future direction for this study could be investigating different scenarios under which multiple waves of epidemic occurs. Another future research idea would be to apply other containment strategies and comparing the results, as well as testing other parameter settings.

## Supporting information

all latex files

## Data Availability

The data that support the findings of this study are available upon request from the corresponding author.

## Appendix 1

The model can be implemented as a classical well-mixed ordinary differential equation system. The governing differential equations are:

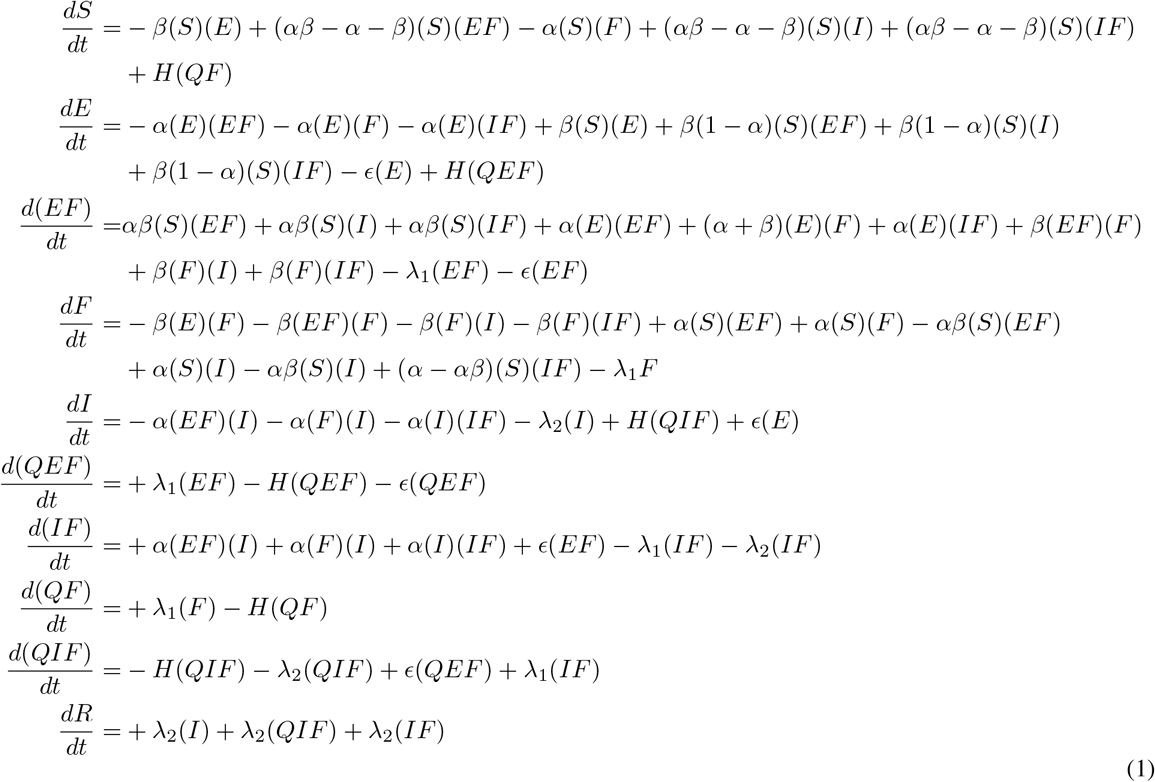

## Appendix 2

The method used to calculate the basic reproduction number is called generation matrix and is described in [18]. With this method, *R*_0_ is defined as spectral radius of the ‘next generation operator’. If the population consists of *n* compartments, in which *m* compartments are infected, we define the vector 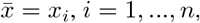, where *x*_*i*_ denotes the number of individuals in the *i*th compartments. Each compartment consists of 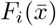 which is the rate of appearance of new infections in compartment *i* and 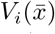 which includes all other transfers of individuals between compartments. The rate of change of each compartment *x*_*i*_ is therefore 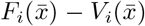. If **G** denotes the next generation matrix, it multiplying is comprised of multiplication of two matrices: **G**_*m*×*m*_ = *FV* ^−1^, where 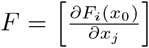 and 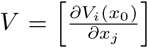. After multiplying matrix *F* by the inverse of matrix *V*, the spectral radius (largest eigenvalue) of the resultant matrix will be *R*_0_. In our model, the infected compartments include E, EF, I, QEF, IF, and QIF compartments. *F* and *V* are then calculated:

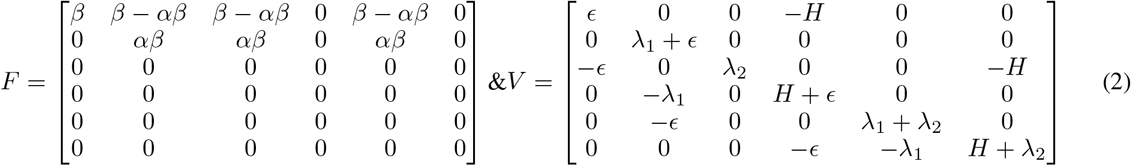

Matrix **G** is easily calculable by calculating *FV* ^−1^, and then the largest eigenvalue of **G** would be the *R*_0_ of the model.

