## Supplementary material for "Investigating dynamics of COVID-19 spread and containment with agent-based modeling": all latex files: bibfile.bib

%% This BibTeX bibliography file was created using BibDesk.
%% http://bibdesk.sourceforge.net/

%% Created for Charlotte Wainwright at 2018-02-12 12:24:13 -0500 


%% Saved with string encoding Unicode (UTF-8) 
@article{giordano2020modelling,
  title={Modelling the COVID-19 epidemic and implementation of population-wide interventions in Italy},
  author={Giordano, Giulia and Blanchini, Franco and Bruno, Raffaele and Colaneri, Patrizio and Di Filippo, Alessandro and Di Matteo, Angela and Colaneri, Marta},
  journal={Nature Medicine},
  pages={1--6},
  year={2020},
  publisher={Nature Publishing Group}
}

@article{sohrabi2020world,
  title={World Health Organization declares global emergency: A review of the 2019 novel coronavirus (COVID-19)},
  author={Sohrabi, Catrin and Alsafi, Zaid and O’Neill, Niamh and Khan, Mehdi and Kerwan, Ahmed and Al-Jabir, Ahmed and Iosifidis, Christos and Agha, Riaz},
  journal={International Journal of Surgery},
  year={2020},
  publisher={Elsevier}
}
@article{heesterbeek2002brief,
  title={A brief history of R 0 and a recipe for its calculation},
  author={Heesterbeek, Johan Andre Peter},
  journal={Acta biotheoretica},
  volume={50},
  number={3},
  pages={189--204},
  year={2002},
  publisher={Springer}
}

@article{pham2020estimating,
  title={On estimating the number of deaths related to Covid-19},
  author={Pham, Hoang},
  journal={Mathematics},
  volume={8},
  number={5},
  pages={655},
  year={2020},
  publisher={Multidisciplinary Digital Publishing Institute}
}

@article{xu2020beware,
  title={Beware of the second wave of COVID-19},
  author={Xu, Shunqing and Li, Yuanyuan},
  journal={The Lancet},
  volume={395},
  number={10233},
  pages={1321--1322},
  year={2020},
  publisher={Elsevier}
}

@article{epstein2008coupled,
  title={Coupled contagion dynamics of fear and disease: mathematical and computational explorations},
  author={Epstein, Joshua M and Parker, Jon and Cummings, Derek and Hammond, Ross A},
  journal={PLoS One},
  volume={3},
  number={12},
  pages={e3955},
  year={2008},
  publisher={Public Library of Science}
}

@book{taleb2007black,
  title={The black swan: The impact of the highly improbable},
  author={Taleb, Nassim Nicholas},
  volume={2},
  year={2007},
  publisher={Random house}
}


@article{lauer2020incubation,
  title={The incubation period of coronavirus disease 2019 (COVID-19) from publicly reported confirmed cases: estimation and application},
  author={Lauer, Stephen A and Grantz, Kyra H and Bi, Qifang and Jones, Forrest K and Zheng, Qulu and Meredith, Hannah R and Azman, Andrew S and Reich, Nicholas G and Lessler, Justin},
  journal={Annals of internal medicine},
  volume={172},
  number={9},
  pages={577--582},
  year={2020},
  publisher={American College of Physicians}
}


@article{thunstrom2020benefits,
  title={The benefits and costs of using social distancing to flatten the curve for COVID-19},
  author={Thunstr{\"o}m, Linda and Newbold, Stephen C and Finnoff, David and Ashworth, Madison and Shogren, Jason F},
  journal={Journal of Benefit-Cost Analysis},
  pages={1--27},
  year={2020},
  publisher={Cambridge University Press}
}


@inproceedings{tisue2004netlogo,
  title={Netlogo: A simple environment for modeling complexity},
  author={Tisue, Seth and Wilensky, Uri},
  booktitle={International conference on complex systems},
  volume={21},
  pages={16--21},
  year={2004},
  organization={Boston, MA}
}

@article{salathe2020covid,
  title={COVID-19 epidemic in Switzerland: on the importance of testing, contact tracing and isolation.},
  author={Salath{\'e}, Marcel and Althaus, Christian L and Neher, Richard and Stringhini, Silvia and Hodcroft, Emma and Fellay, Jacques and Zwahlen, Marcel and Senti, Gabriela and Battegay, Manuel and Wilder-Smith, Annelies and others},
  journal={Swiss medical weekly},
  volume={150},
  number={11-12},
  pages={w20225},
  year={2020},
  publisher={EMH Swiss Medical Publishers, Ltd.}
}

@misc{cohen2020countries,
  title={Countries test tactics in ‘war’against COVID-19},
  author={Cohen, Jon and Kupferschmidt, Kai},
  year={2020},
  publisher={American Association for the Advancement of Science}
}


@article{saez2020effectiveness,
  title={Effectiveness of the measures to flatten the epidemic curve of COVID-19. The case of Spain},
  author={Saez, Marc and Tobias, Aurelio and Varga, Diego and Barcel{\'o}, Maria Ant{\`o}nia},
  journal={Science of the Total Environment},
  pages={138761},
  year={2020},
  publisher={Elsevier}
}


@article{moss2020modelling,
  title={Modelling the impact of COVID-19 in Australia to inform transmission reducing measures and health system preparedness},
  author={Moss, Robert and Wood, James and Brown, Damien and Shearer, Freya and Black, Andrew J and Cheng, Allen and McCaw, James M and McVernon, Jodie},
  journal={medRxiv},
  year={2020},
  publisher={Cold Spring Harbor Laboratory Press}
}


@article{verity2020estimates,
  title={Estimates of the severity of coronavirus disease 2019: a model-based analysis},
  author={Verity, Robert and Okell, Lucy C and Dorigatti, Ilaria and Winskill, Peter and Whittaker, Charles and Imai, Natsuko and Cuomo-Dannenburg, Gina and Thompson, Hayley and Walker, Patrick GT and Fu, Han and others},
  journal={The Lancet infectious diseases},
  year={2020},
  publisher={Elsevier}
}

@article{kermack1927contribution,
  title={A contribution to the mathematical theory of epidemics},
  author={Kermack, William Ogilvy and McKendrick, Anderson G},
  journal={Proceedings of the royal society of london. Series A, Containing papers of a mathematical and physical character},
  volume={115},
  number={772},
  pages={700--721},
  year={1927},
  publisher={The Royal Society London}
}


@article{lu2003analysis,
  title={Analysis of an SI epidemic model with nonlinear transmission and stage structure},
  author={Lu, Zhonghua and Gao, Shujing and Chen, Lansun},
  journal={Acta Mathematica Scientia},
  volume={23},
  number={4},
  pages={440--446},
  year={2003},
  publisher={Elsevier}
}


@article{mishra2010seiqrs,
  title={SEIQRS model for the transmission of malicious objects in computer network},
  author={Mishra, Bimal Kumar and Jha, Navnit},
  journal={Applied Mathematical Modelling},
  volume={34},
  number={3},
  pages={710--715},
  year={2010},
  publisher={Elsevier}
}


@book{daley2001epidemic,
  title={Epidemic modelling: an introduction},
  author={Daley, Daryl J and Gani, Joe},
  volume={15},
  year={2001},
  publisher={Cambridge University Press}
}

@article{kelly2013selecting,
  title={Selecting among five common modelling approaches for integrated environmental assessment and management},
  author={Kelly, Rebecca A and Jakeman, Anthony J and Barreteau, Olivier and Borsuk, Mark E and ElSawah, Sondoss and Hamilton, Serena H and Henriksen, Hans J{\o}rgen and Kuikka, Sakari and Maier, Holger R and Rizzoli, Andrea Emilio and others},
  journal={Environmental modelling \& software},
  volume={47},
  pages={159--181},
  year={2013},
  publisher={Elsevier}
}

@article{hackl2019epidemic,
  title={Epidemic spreading in urban areas using agent-based transportation models},
  author={Hackl, J{\"u}rgen and Dubernet, Thibaut},
  journal={Future Internet},
  volume={11},
  number={4},
  pages={92},
  year={2019},
  publisher={Multidisciplinary Digital Publishing Institute}
}


@article{epstein2009modelling,
  title={Modelling to contain pandemics},
  author={Epstein, Joshua M},
  journal={Nature},
  volume={460},
  number={7256},
  pages={687--687},
  year={2009},
  publisher={Nature Publishing Group}
}


@article{johnson2002updating,
  title={Updating the accounts: global mortality of the 1918-1920" Spanish" influenza pandemic},
  author={Johnson, Niall PAS and Mueller, Juergen},
  journal={Bulletin of the History of Medicine},
  pages={105--115},
  year={2002},
  publisher={JSTOR}
}

----------------------------------
%%contribution
@article{tyson2020timing,
  title={The Timing and Nature of Behavioural Responses Affect the Course of an Epidemic},
  author={Tyson, Rebecca C and Hamilton, Stephanie D and Lo, Aboubakr S and Baumgaertner, Bert O and Krone, Stephen M},
  journal={Bulletin of Mathematical Biology},
  volume={82},
  number={1},
  pages={14},
  year={2020},
  publisher={Springer}
}


@article{chang2020modelling,
  title={Modelling transmission and control of the COVID-19 pandemic in Australia},
  author={Chang, Sheryl L and Harding, Nathan and Zachreson, Cameron and Cliff, Oliver M and Prokopenko, Mikhail},
  journal={arXiv preprint arXiv:2003.10218},
  year={2020}
}


@article{gros2020containment,
  title={Containment efficiency and control strategies for the Corona pandemic costs},
  author={Gros, Claudius and Valenti, Roser and Schneider, Lukas and Valenti, Kilian and Gros, Daniel},
  journal={arXiv preprint arXiv:2004.00493},
  volume={1},
  year={2020}
}

@article{cuevas2020agent,
  title={An agent-based model to evaluate the COVID-19 transmission risks in facilities},
  author={Cuevas, Erik},
  journal={Computers in Biology and Medicine},
  pages={103827},
  year={2020},
  publisher={Elsevier}
}


@article{silva2020covid,
  title={COVID-ABS: An Agent-Based Model of COVID-19 Epidemic to Simulate Health and Economic Effects of Social Distancing Interventions},
  author={Silva, Petr{\^o}nio CL and Batista, Paulo VC and Lima, H{\'e}lder S and Alves, Marcos A and Guimar{\~a}es, Frederico G and Silva, Rodrigo CP},
  journal={Chaos, Solitons \& Fractals},
  pages={110088},
  year={2020},
  publisher={Elsevier}
}


@article{azizi2020epidemics,
  title={Epidemics on networks: Reducing disease transmission using health emergency declarations and peer communication},
  author={Azizi, Asma and Montalvo, Cesar and Espinoza, Baltazar and Kang, Yun and Castillo-Chavez, Carlos},
  journal={Infectious Disease Modelling},
  volume={5},
  pages={12--22},
  year={2020},
  publisher={Elsevier}
}


@article{sadeghi2020universal,
  title={Universal features of epidemic models under social distancing guidelines},
  author={Sadeghi, Mahdiar and Greene, James and Sontag, Eduardo},
  journal={bioRxiv},
  year={2020},
  publisher={Cold Spring Harbor Laboratory}
}


@article{o2010agent,
  title={Agent-based modeling of the spread of the 1918--1919 flu in three Canadian fur trading communities},
  author={O'neil, Caroline A and Sattenspiel, Lisa},
  journal={American Journal of Human Biology},
  volume={22},
  number={6},
  pages={757--767},
  year={2010},
  publisher={Wiley Online Library}
}

@article{carpenter2009design,
  title={The design and use of an agent-based model to simulate the 1918 influenza epidemic at Norway House, Manitoba},
  author={Carpenter, Connie and Sattenspiel, Lisa},
  journal={American Journal of Human Biology: The Official Journal of the Human Biology Association},
  volume={21},
  number={3},
  pages={290--300},
  year={2009},
  publisher={Wiley Online Library}
}

@article{dimka2014applications,
  title={Applications of Agent-Based Modelling Techniques to Studies of Historical Epidemics: The 1918 Flu in Newfoundland and Labrador},
  author={Dimka, Jessica and Orbann, Carolyn and Sattenspiel, Lisa},
  journal={Journal of the Canadian Historical Association/Revue de la Soci{\'e}t{\'e} historique du Canada},
  volume={25},
  number={2},
  pages={265--296},
  year={2014},
  publisher={The Canadian Historical Association/La Soci{\'e}t{\'e} historique du Canada}
}


@article{crooks2014agent,
  title={An agent-based modeling approach applied to the spread of cholera},
  author={Crooks, Andrew T and Hailegiorgis, Atesmachew B},
  journal={Environmental Modelling \& Software},
  volume={62},
  pages={164--177},
  year={2014},
  publisher={Elsevier}
}


@incollection{khalil2012agent,
  title={An agent-based modeling for pandemic influenza in Egypt},
  author={Khalil, Khaled M and Abdel-Aziz, M and Nazmy, Taymour T and Salem, Abdel-Badeeh M},
  booktitle={Handbook on Decision Making},
  pages={205--218},
  year={2012},
  publisher={Springer}
}

@article{perez2009agent,
  title={An agent-based approach for modeling dynamics of contagious disease spread},
  author={Perez, Liliana and Dragicevic, Suzana},
  journal={International journal of health geographics},
  volume={8},
  number={1},
  pages={50},
  year={2009},
  publisher={Springer}
}

@article{siettos2015modeling,
  title={Modeling the 2014 ebola virus epidemic--agent-based simulations, temporal analysis and future predictions for liberia and sierra leone},
  author={Siettos, Constantinos and Anastassopoulou, Cleo and Russo, Lucia and Grigoras, Christos and Mylonakis, Eleftherios},
  journal={PLoS currents},
  volume={7},
  year={2015},
  publisher={Public Library of Science}
}


@article{heffernan2005perspectives,
  title={Perspectives on the basic reproductive ratio},
  author={Heffernan, Jane M and Smith, Robert J and Wahl, Lindi M},
  journal={Journal of the Royal Society Interface},
  volume={2},
  number={4},
  pages={281--293},
  year={2005},
  publisher={The Royal Society London}
}

@article{van2017reproduction,
  title={Reproduction numbers of infectious disease models},
  author={van den Driessche, Pauline},
  journal={Infectious Disease Modelling},
  volume={2},
  number={3},
  pages={288--303},
  year={2017},
  publisher={Elsevier}
}

@article{crooks2013gis,
  title={GIS and agent-based models for humanitarian assistance},
  author={Crooks, Andrew T and Wise, Sarah},
  journal={Computers, Environment and Urban Systems},
  volume={41},
  pages={100--111},
  year={2013},
  publisher={Elsevier}
}

@inproceedings{wang2010use,
  title={Use of GIS and agent-based modeling to simulate the spread of influenza},
  author={Wang, Jiasheng and Xiong, Jianhong and Yang, Kun and Peng, Shuangyun and Xu, Quanli},
  booktitle={2010 18th International Conference on Geoinformatics},
  pages={1--6},
  year={2010},
  organization={IEEE}
}


@inproceedings{cui2009exploring,
  title={Exploring HIV/AIDS epidemic complex network of IDU using ABM and GIS},
  author={Cui, Qingxiong and Wang, Jun and Tan, Junxiang and Li, Jiangrong and Yang, Kun},
  booktitle={2009 Chinese Control and Decision Conference},
  pages={1090--1095},
  year={2009},
  organization={IEEE}
}


@article{hale2020variation,
  title={Variation in government responses to COVID-19},
  author={Hale, Thomas and Petherick, Anna and Phillips, Toby and Webster, Samuel},
  journal={Blavatnik school of government working paper},
  volume={31},
  year={2020}
}


@article{kramer2014experimental,
  title={Experimental evidence of massive-scale emotional contagion through social networks},
  author={Kramer, Adam DI and Guillory, Jamie E and Hancock, Jeffrey T},
  journal={Proceedings of the National Academy of Sciences},
  volume={111},
  number={24},
  pages={8788--8790},
  year={2014},
  publisher={National Acad Sciences}
}
