## Supplementary material for "Investigating dynamics of COVID-19 spread and containment with agent-based modeling": all latex files: README.md

## Description:

The project hosts an aesthetic an simple LaTeX style suitable for "preprint" publications such as arXiv and bio-arXiv, etc. 
It is based on the [**nips_2018.sty**](https://media.nips.cc/Conferences/NIPS2018/Styles/nips_2018.sty) style.

This styling maintains the esthetic of NIPS but adding and changing features to make it (IMO) even better and nore suitable for preprints.
The result looks fairly different from NIPS style so that readers won't get confused to think that the preprint was published in NIPS. 

### Why NIPS? 
Because the NIPS styling is a comfortable single column format that is very esthetic and convenient for reading.

## Usage:
1. Use Document class **article**. 
2. Copy **arxiv.sty** to the folder containing your tex file.
3. add `\usepackage{arxiv}` after `\documentclass{article}`.
4. The only packages used in the style file are **geometry** and **fancyheader**. Do not reimport them.

See **template.tex** 

## Project files:
1. **arxiv.sty** - the style file.
2. **template.tex** - a sample template that uses the **arxiv style**.
3. **references.bib** - the bibliography source file for template.tex.
4. **template.pdf** - a sample output of the template file that demonstrated the design provided by the arxiv style.


## Handling References when submitting to arXiv.org
The most convenient way to manage references is using an external BibTeX file and pointing to it from the main file. 
However, this requires running the [bibtex](http://www.bibtex.org/) tool to "compile" the `.bib` file and create `.bbl` file containing "bibitems" that can be directly inserted in the main tex file. 
However, unfortunately the arXiv Tex environment ([Tex Live](https://www.tug.org/texlive/)) do not do that. 
So easiest way when submitting to arXiv is to create a single self-contained .tex file that contains the references.
This can be done by running the BibTeX command on your machine and insert the content of the generated `.bbl` file into the `.tex` file and commenting out the `\bibliography{references}` that point to the external references file.

Below are the commands that should be run in the project folder:
1. Run `$ latex template`
2. Run `$ bibtex template`
3. A `template.bbl` file will be generated (make sure it is there)
4. Copy the `template.bbl` file content to `template.tex` into the `\begin{thebibliography}` command.
5. Comment out the `\bibliography{references}` command in `template.tex`.
6. You ready to submit to arXiv.org.


## General Notes:
1. For help, comments, praises, bug reporting or change requests, you can contact the author at: kourgeorge/at/gmail.com.
2. You can use, redistribute and do whatever with this project, however, the author takes no responsibility on whatever usage of this project.
3. If you start another project based on this project, it would be nice to mention/link to this project.
4. You are very welcome to contribute to this project.
5. A good looking 2 column template can be found in https://github.com/brenhinkeller/preprint-template.tex.
