## Supplementary figures and images for "Investigating dynamics of COVID-19 spread and containment with agent-based modeling"

### SEIR-edited.PNG

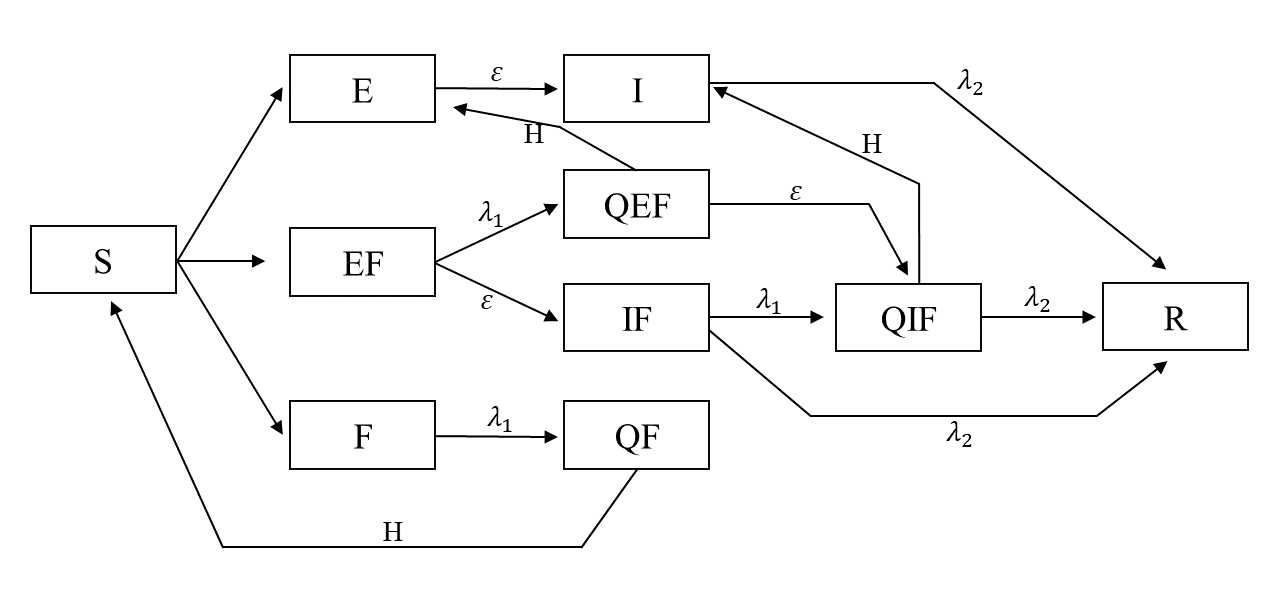

### Test-With-Tracking.png

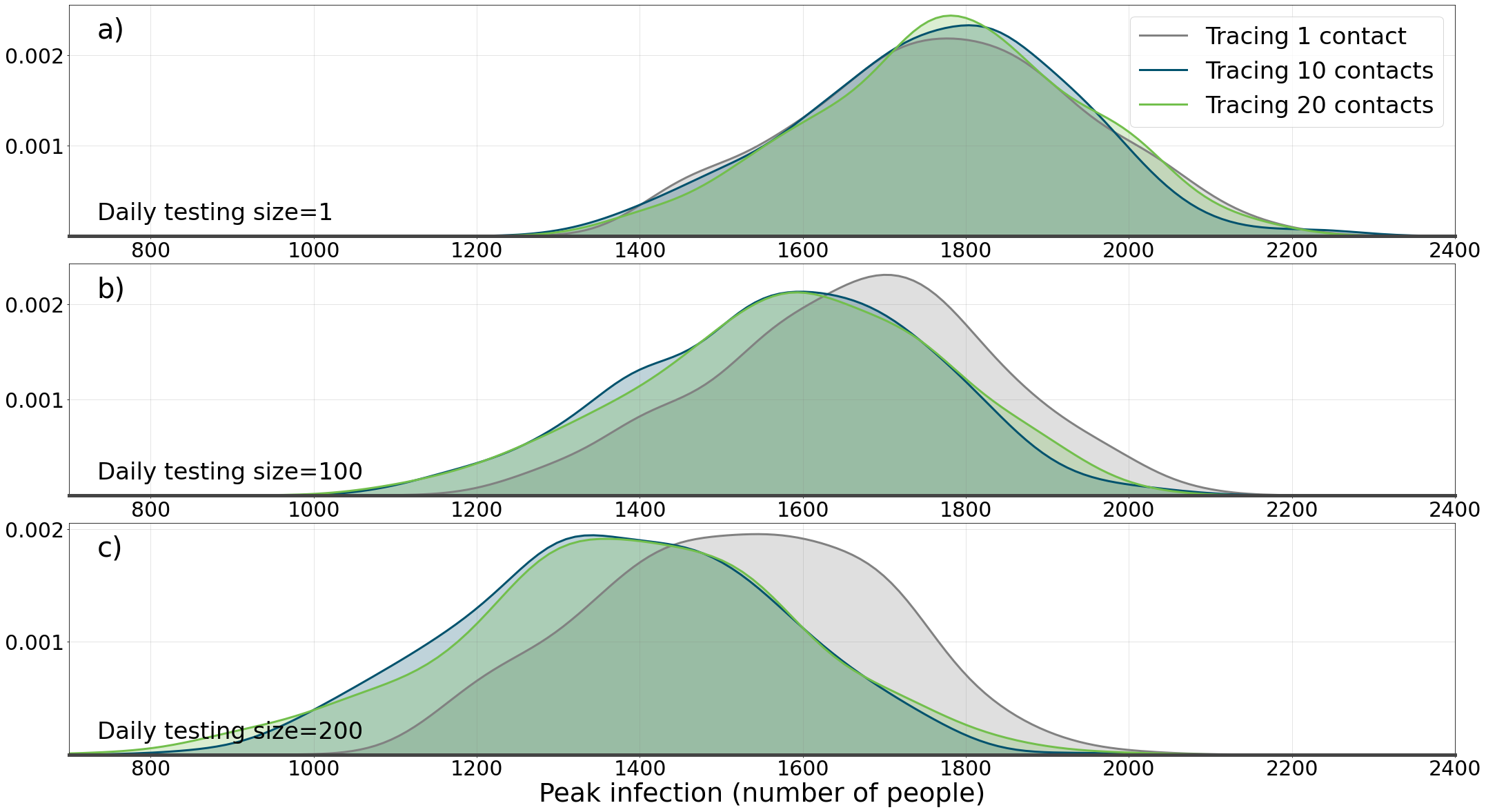

### twowaves2.png

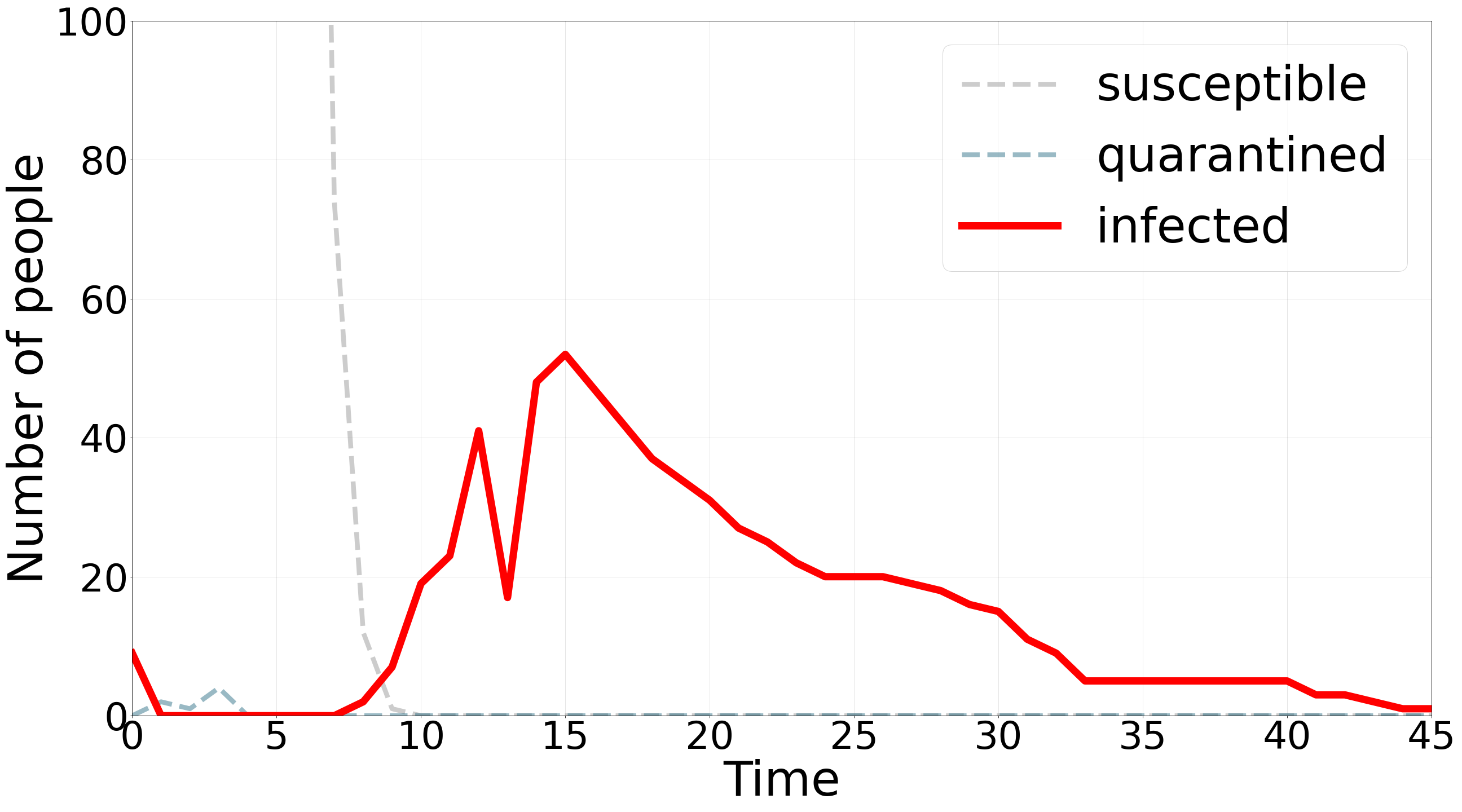

### twowaves.png

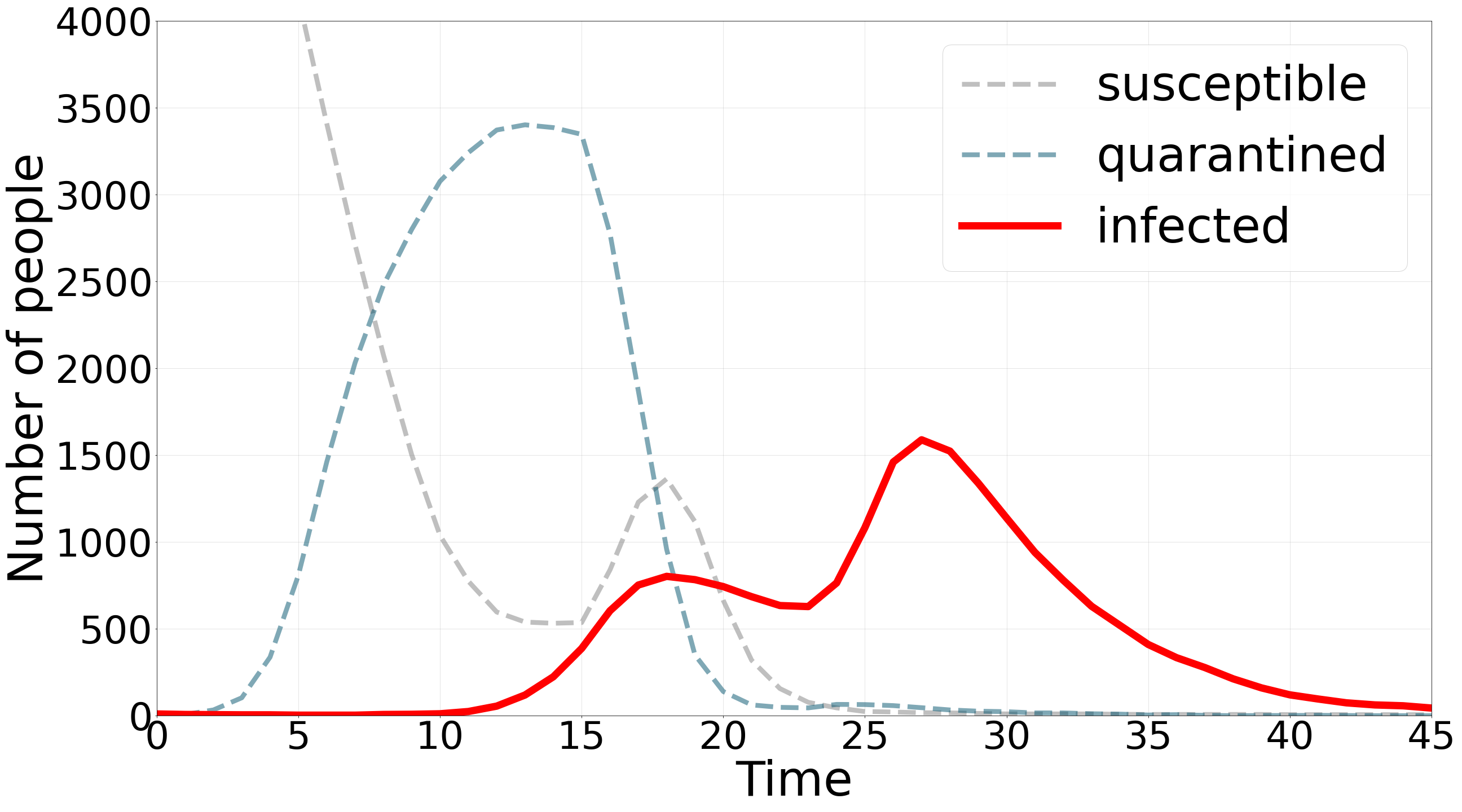

### withandwithout-tracking.png

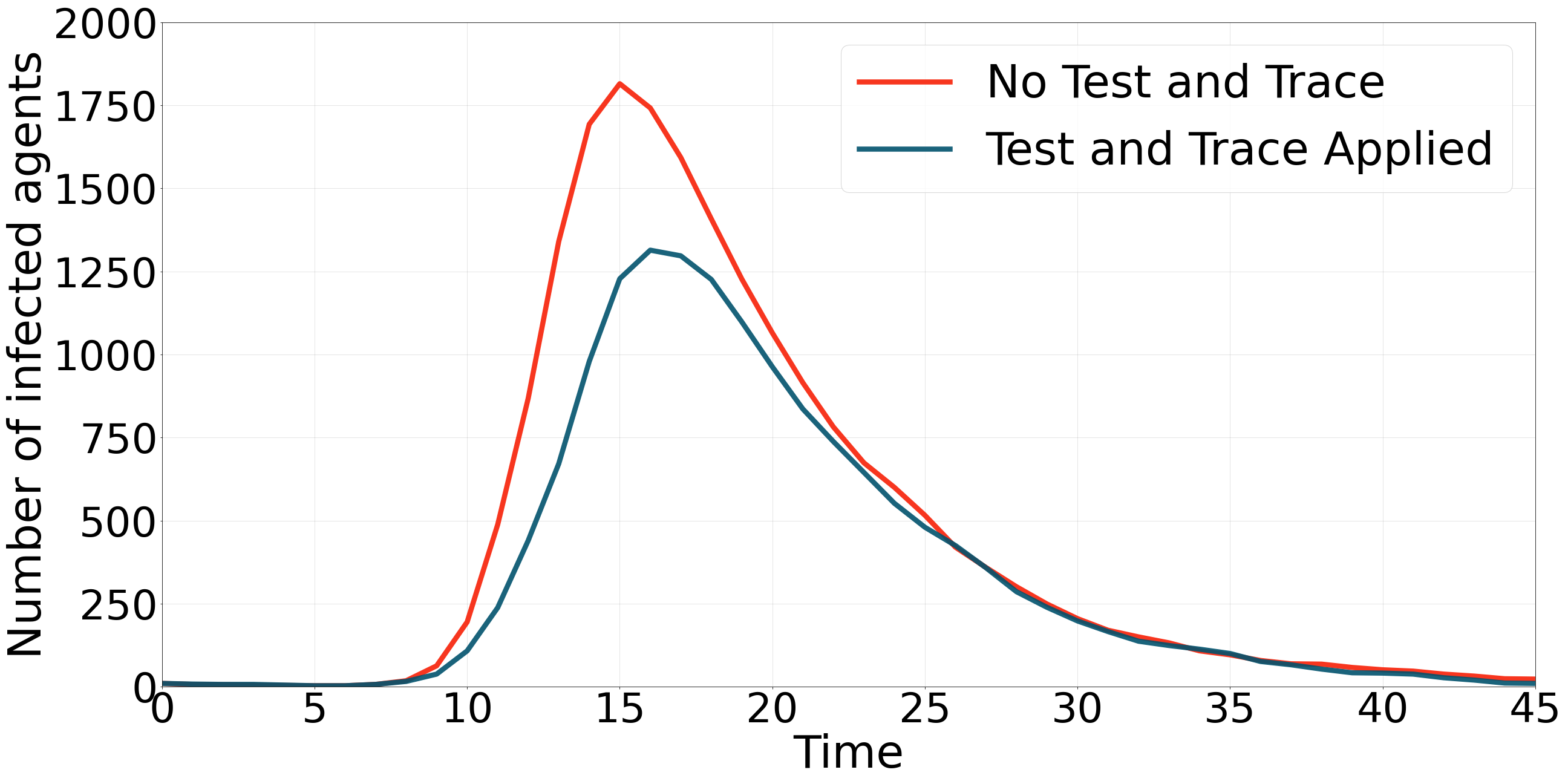

### withandwithout-zoning.png

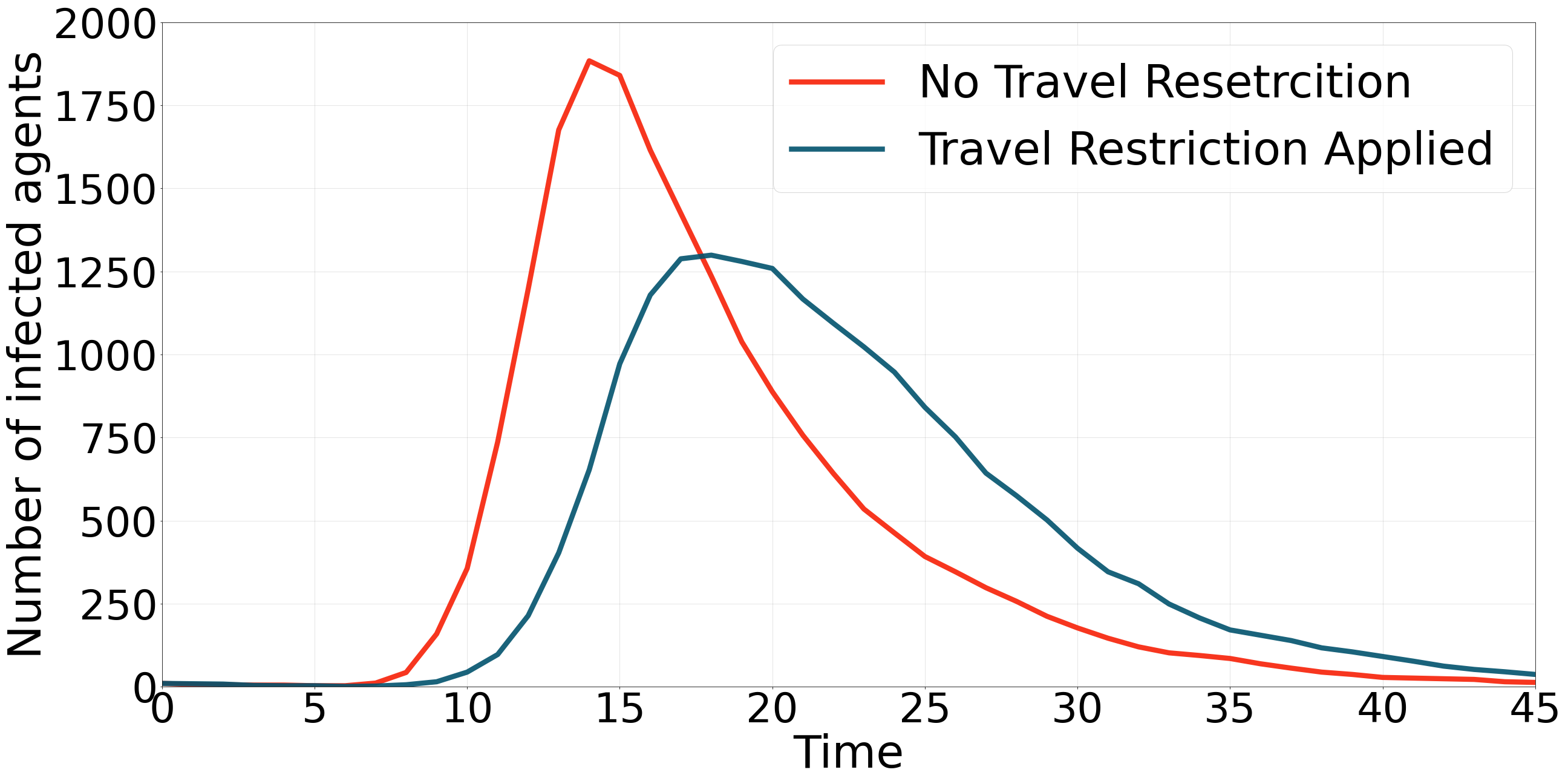

### zoning.png

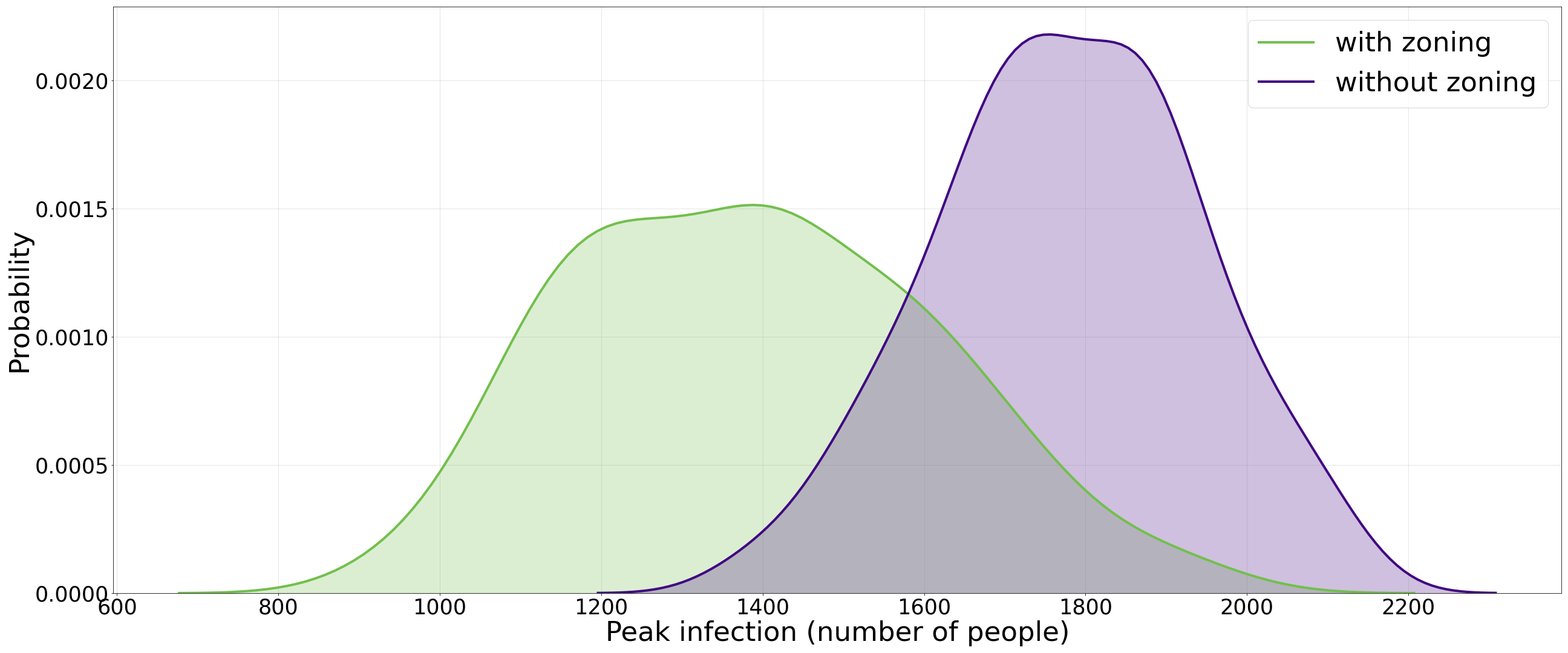
